# System Integrated Digital Empowerment and Rehabilitation to promote patient Activation and well-Being (SIDERA^B): Protocol for a Randomized Crossover Trial on Effectiveness and Implementation

**DOI:** 10.1101/2022.04.19.22274012

**Authors:** Federica Rossetto, Francesca Borgnis, Valeria Blasi, Paolo Innocente Banfi, Monica Tavanelli, Olivia Realdon, Fabrizia Mantovani, Emanuela Foglia, Elisabetta Garagiola, Davide Croce, Marco Nalin, Francesca Baglio

## Abstract

**CONTEXT:** the current increasing demand for rehabilitation among people with Non-Communicable Diseases (NCDs) requires the identification of home-based digital solutions alternative to conventional in-clinic interventions.

**OBJECTIVE:** this protocol proposes to test the effectiveness of an individualized telerehabilitation platform (SIDERA^B), with respect to the traditional face-to-face rehabilitation, in ensuring the continuity of care in patients with NCDs.

**DESIGN, SETTING, AND SUBJECTS:** this randomized, single-blind, controlled two-period crossover trial will involve about 150 outpatients with NCDs (N=40 with Chronic Heart Failure – CHF, N=60 with Chronic Obstructive Pulmonary Disease – COPD, and N=50 with Parkinson’s Disease – PD) from the rehabilitation units of IRCCS Fondazione Don Carlo Gnocchi of Milan. Each participant will experience, consequently, two different types of interventions: rehabilitation with the SIDERA^B system (SIDERA^B – S), which allow for both tele-rehabilitation activities and tele-monitoring of vital parameters, and rehabilitation as usual (Usual Care – U) including a manual of rehabilitative exercises and self-monitoring of vital parameters.

**INTERVENTIONS:** subjects will be randomly assigned to one of the two specified sequences of interventions: U/S/U (the USU group), and S/U (the SU group). Both groups will be assessed at the baseline (T1), after the first intervention (T2), and after the second intervention (T3), with a follow-up evaluation (T4) scheduled only for the USU group.

**MAIN OUTCOME MEASURES:** a multifaceted evaluation including quality of life and clinical/functional measures will be conducted at each time-point of assessment. The primary outcome measures will be 1) change in activation of patients measured by the Patient Activation Measure scale, and 2) change in subject’s level of activity and participation measured by the WHO Disability Assessment Schedule 2.0.

**CONCLUSION:** SIDERA^B could represent a promising innovative digital solution able to support the ongoing migration of rehabilitation care from the clinic to the patient’s home, for the optimal long-term management of NCDs.

**Trial registration:** The SIDERA^B trial was registered in the clinicaltrials.gov database (identifier NCT04041193) on August 1, 2019.

## Background

As people live longer, but also with a greater number of long-term conditions (also called “non-communicable diseases” - NCDs), there is an unavoidable increase in the demand for healthcare (Topol, 2019a). Such demographic and health evolution is leading to rapid growth in the number of people experiencing disability or declines in daily functioning. The *Global Burden of Diseases, Injuries, and Risk Factors Study* (GBD 2019, Vos et al., 2020) estimates that, since 1990, there has been a pronounced shift towards a greater proportion of burden due to YLDs (*Years of Life lived with Disability* - YLD) from NCDs. In 2019, NCDs and injury YLDs contributed to more than half of all disease burdens in 11 countries. Only in Europe, it is estimated that chronic diseases are responsible for 86% of all deaths and healthcare costs are evaluated for an impact equal to 700 billion euros per year (Topol, 2019a). Diseases such as heart failure, diabetes, obesity and respiratory failure affect about 80% of people over the age of 65 and by 2060 this number is expected to increase from 88 to 152 million (Italian National Plan of Chronicity, 2016). In this era of global health changes, a radical transformation of the healthcare system requiring the identification of alternatives to the hospital is essential. Furthermore, unprecedented new challenges to patient care have been determined by the COVID-19 pandemic, including difficulties accessing routine treatments, for people with NCDs.

Current technological advances in digital medicine are driving those changes. Digital healthcare technologies, defined as genomics, digital medicine, artificial intelligence and robotics, are seen as new means of addressing the big healthcare challenges of the 21st century (Topol, 2019a). Telemedicine is an example of a technology already in progress for the management of chronic conditions, even with partial and discontinuous adoption across national healthcare systems. It involves the delivery of remote clinical care using telecommunication and information technologies, complying with the standards required by face-to-face interventions (Topol, 2019b; Dorsey et al., 2016). The main goal is to build an inter-professional, community-based network to support older adults staying in their homes, improving both the self-management and the efficiency of home-care providers by enhancing the potential of technology (Matthew-Maich et al., 2016). Moreover, the growing appeal of mobile solutions such as Apps, linked to sensors and wearable devices for remote vital-parameters monitoring, is a reality: these solutions began to be prescribed as digital therapeutics for highly customized self-monitoring and self-management of health conditions.

In this framework, telerehabilitation meets the need to make rehabilitation a powerful, widespread service, ensuring accessibility, quality of care and high levels of patient engagement in the continuum of care. Also, telerehabilitation constitutes an innovative way to provide long-lasting, technology-enabled rehabilitation care outside the hospital settings through a “double-loop” communication between the clinic and the patient’s home (Di Tella et al., 2020; Isernia et al., 2020, 2019), a crucial communication component that enables both remote monitoring of patient performance and responding with appropriate feedback (Di Tella et al., 2020; Alaimo et al., 2021).

This type of communication represents a crucial requirement, especially for asynchronous models of telerehabilitation (namely, when patient and therapist do not interact in real-time), in which a digital platform enables the “assessment, monitoring and feedback” loop in a complex ecosystem. The integration of digital contents constitutes an additional key aspect of telerehabilitation, providing the clinician with real-time monitoring data (adherence/performance) and enhancing patient’s participation and engagement in their individualized healthcare processes (Matamala-Gomez et al., 2020).

In the last years, telerehabilitation has proved to be a promising option for the successful management of NCDs in terms of healthcare delivery, increased compliance to treatment, improved health outcomes/quality of life, and reduced costs (Velayati et al., 2020; Peretti et al., 2017). However, large-scale randomized controlled trials demonstrating the clinical-effectiveness of these solutions are still limited.

The main aim of the proposed randomized, double-blind, crossover trial is to test the effectiveness of an individualized, home-based telerehabilitation program, with respect to the traditional face-to-face rehabilitation, in ensuring the double-loop communication of the continuity of care among patients with Chronic Heart Failure (CHF), Chronic Obstructive Pulmonary Disease (COPD) and Parkinson’s Disease (PD). In particular, the “System Integrated Digital Empowerment and Rehabilitation to promote patient Activation and well-Being” (SIDERA^B) will be tested. We will investigate after-treatment changes in the “Patient-Relevant Structural and Procedural Effects” and in outcomes referring to the “Medical Benefits” field comparing patients in the “SU group”, in which they will experience first the SIDERA^B system and then the Usual Care intervention, and the “USU group”, in which subjects will attempt the Usual Care intervention followed by the SIDERA^B system, and by an additional Usual Care intervention.

## Methods

The protocol of the study has been prepared as outlined in the “Standard Protocol Items: Recommendations for Interventional Trials” (SPIRIT) guidelines (Figure 1). The study will be conducted according to the Declaration of Helsinki, the principles of Good Clinical Practice, and in accordance with local legislation in participating countries.

**FIGURE 1.:**
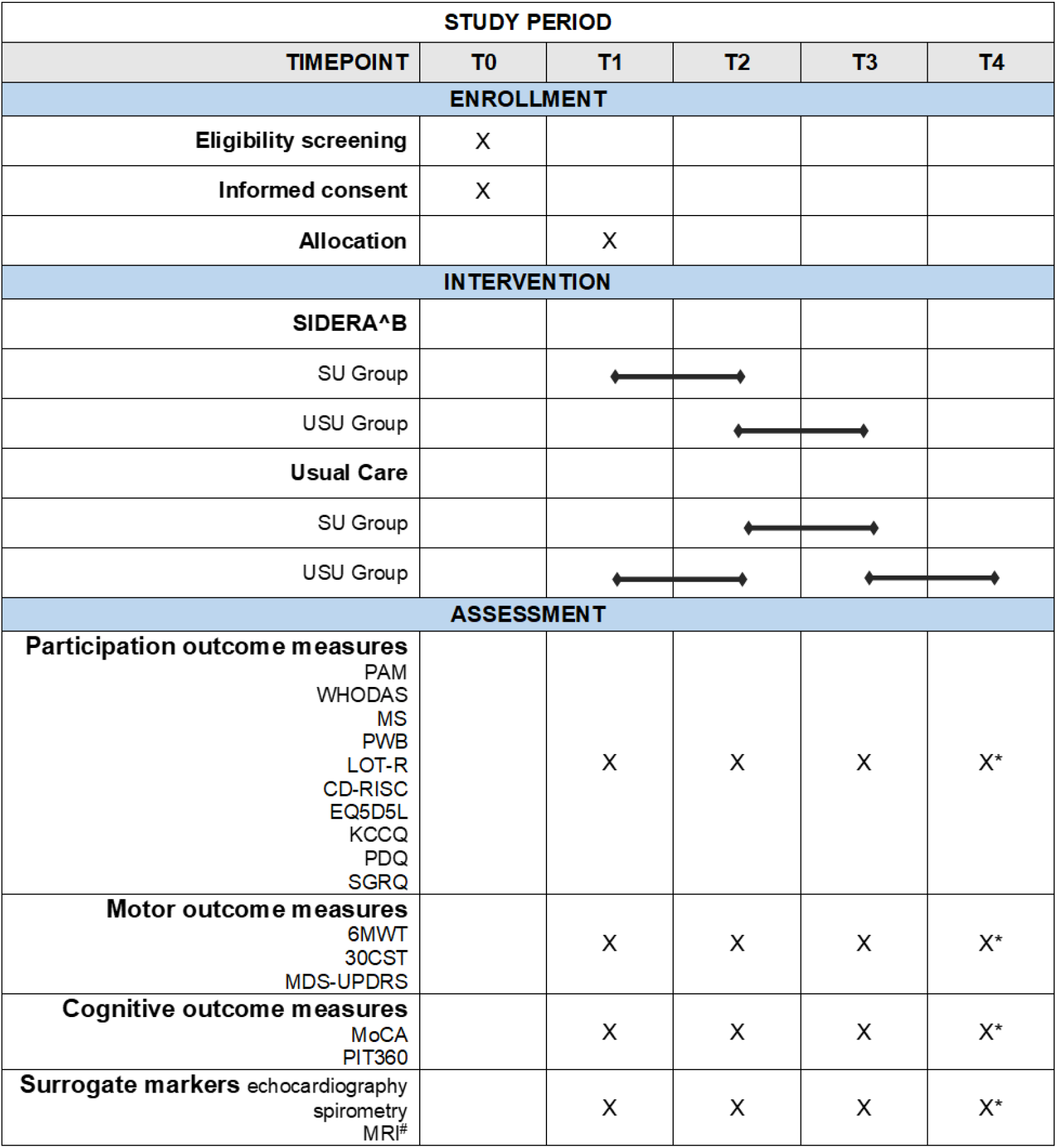
SPIRIT figure for the schedule of enrolment, interventions, and assessments in a crossover study design. T0= pre-intervention phase; T1=baseline assessment; T2 = assessment after the first intervention; T3 = assessment after the second intervention; T4 = follow-up evaluation. S= SIDERA^B; U= Usual Care. QoL=Quality of Life. 30CST: 30 Second Sit to Stand Test; 6MWT: 6’ Minute Walking Test; CD-RISC: Connor-Davidson Resilience Scale; EF: Ejection Fraction; EQ5D5L: EuroQol five dimensions (5D) and five levels (5L); FEV1%: Forced expiratory volume in the 1st second; KCCQ: Kansas City Cardiomyopathy Questionnaire; LOT-R: Life Orientation Test-Revised; MDS-UPDRS: Movement Disorder Society - Unified Parkinson’s Disease Rating Scale; MoCA: Montreal Cognitive Assessment test; MRI: Magnetic Resonance Imaging; MS: Mutuality Scale; PAM13: Patient Activation Measure scale; PANAS: Positive-Negative Affect Schedule; PDQ: Parkinsons’ Disease Questionnaire; PIT360: Picture Interpretation test 360; PWB: Psychological well-being scale; SGRQ: St George’s Respiratory Questionnaire; WHODAS 2.0: WHO Disability Assessment Schedule 2.0. #Facultative *Administered only to the USU group

### Trial design and setting

This study is designed as a single-blinded, randomized, cross-over trial involving outpatients from rehabilitation units of IRCCS Fondazione Don Carlo Gnocchi (Milan, Italy). Each participant will experience, consequently, two different types of interventions: rehabilitation with the SIDERA^B system (SIDERA^B – S) and rehabilitation as usual (Usual Care – U).

After being recruited, subjects will be randomly assigned to one of the two pre-specified sequences of interventions: U/S/U (the USU group), and S/U (the SU group). The duration of study participation will vary depending on the group allocation: the USU group will include four time-point of evaluation (T1 = baseline assessment; T2 = assessment after the first intervention; T3 = assessment after the second intervention; T4 = follow-up evaluation), while three time-point of assessment will be scheduled for subjects in the SU group (T1 = baseline assessment; T2 = assessment after the first intervention; T3 = assessment after the second intervention). The rationale underlying the differences in the number of time-point evaluations between the two groups is the effort to reduce the commitment of patients and their caregivers while taking into consideration the follow-up effects of treatment. The trial work plan is shown in Figure 2.

**FIGURE 2.:**
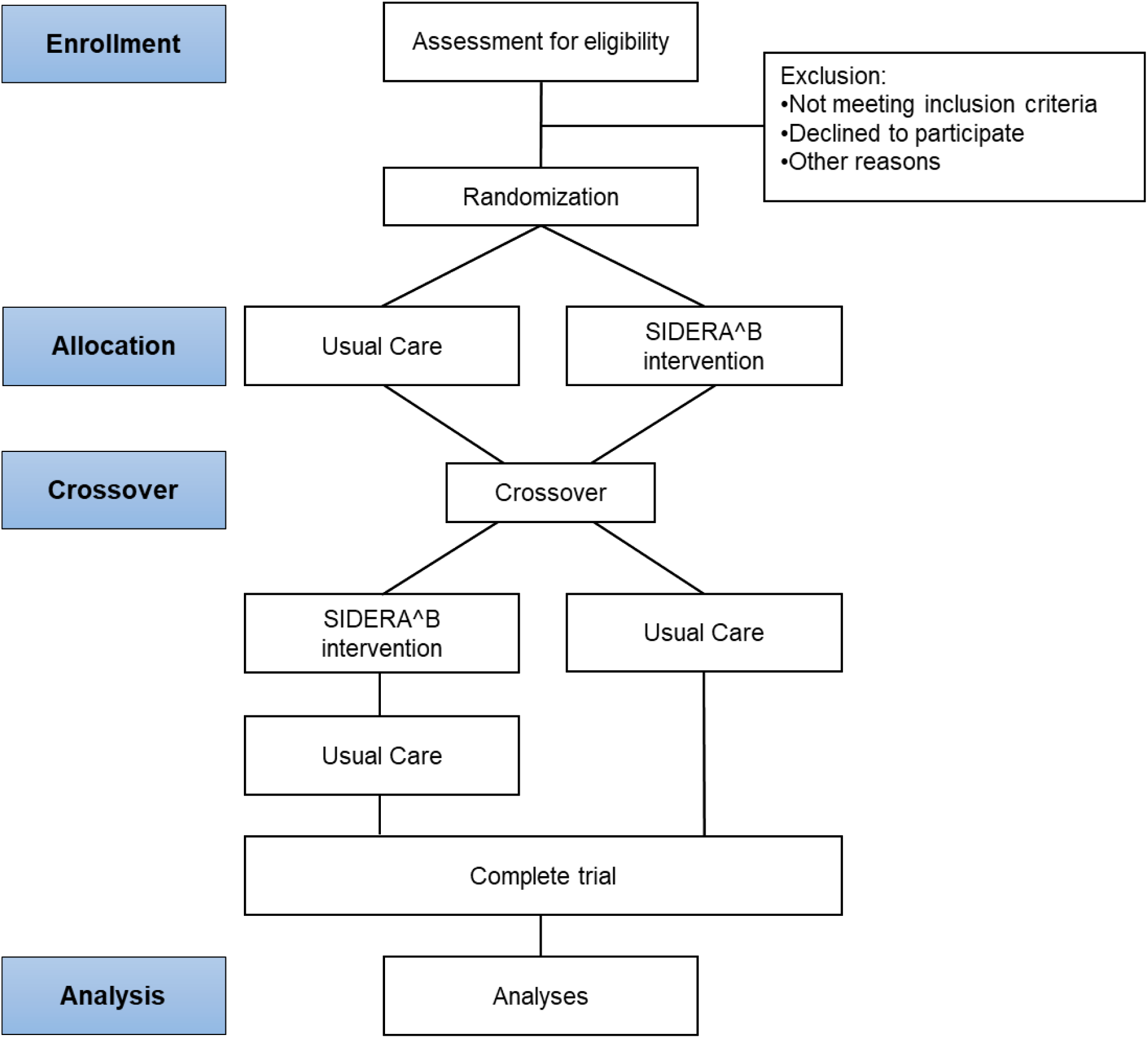
The trial work plan.

### Sample size

The sample size was computed using the *G* Power 3* software (Faul et al., 2007; 2009) according to previous studies (Anderson et al., 2018; Solomon et al., 2012). Under the assumption of normal distribution of the outcome scores, with an estimate of a 5-point difference on a 100-point scale in patient activation scores (Anderson et al., 2018) between pre - and post-intervention measurement, and assuming a standard deviation of 11 for both arms, approximately 60 patients are needed for each arm to obtain a statistical power of 80% (two-sided Type I error rate of 0.05) based on a 1:1 treatment allocation. After considering a dropout rate of approximately 25%, a total of 150 patients with 75 patients per arm will be needed for this trial.

### Study population, recruitment and randomization

According to the sample size calculation, the SIDERA^B trial has a target enrollment of 150 outpatients with the diagnosis of Chronic Heart Failure – CHF (N=40), Chronic Obstructive Pulmonary Disease – COPD (N=60) and Parkinson’s Disease – PD (N=50). Eligible patients who meet all inclusion criteria (see the paragraph below) will be randomized using a web-based allocation concealment through a computer-based algorithm created by an independent statistician. Randomization will be stratified according to NCDs. The trial intervention will not be blinded for clinicians or patients due to its nature. Conversely, clinical endpoints and data collection from clinical/psychological questionnaires will be blinded for examiners/assessors. The statistician conducting the data analysis will be masked for the group allocation.

### Inclusion and exclusion criteria

Inclusion criteria for all participants will be:

1. age between 18 and 85 years (adult and older adult);
2. agreement to participate with the signature of the informed consent form;
3. availability of a caregiver/study partner, who agrees to support the participant through the SIDERA^B trial;
4. living in one’s own home;
5. clinical diagnosis of CHF according to European Society of Cardiology guidelines (Ponikowski et al., 2016) with Functional *New York Heart Association* (NYHA) class II and III and Etiology of primitive or post-ischemic CHF OR clinical diagnosis of COPD according to the American Thoracic Society (ATS) and the European Respiratory Society (ERS) criteria (Brusasco et al., 2005) with the presence of airway limitation according to ATS/ERS and GOLD criteria (Global Initiative for Chronic Obstructive Lung Disease, Report 2018) between 1B and 4C OR clinical diagnosis of PD according to the Movement Disorder Society (MDS) criteria (Postuma et al., 2015), and disease staging between 1.5 and 3 on the Hoehn & Yahr scale (Goetz et al., 2004).

Exclusion criteria will be:

1. presence of comorbidities that might prevent patients from undertaking a safe home program or determining clinical instability (i.e., severe orthopedic or severe cognitive deficits);
2. presence of risk conditions for safety under strain in patients with CHF (i.e., severe symptomatic aortic stenosis, evidence of ischemia due to minimal/low-intensity efforts); CHF Etiology other than primitive or post-ischemic;
3. severe COPD (GOLD 4 / D class) or mild symptoms which do not need rehabilitation treatments (GOLD 1 / A class); overlapping between COPD and other respiratory diseases or not in treatment or in the absence of good ventilatory compensation in the last 6 months; COPD with global respiratory failure with PaCO2> 55 mmHg;
4. overlapping between PD and other neurological pathologies or with severe psychiatric complications; pathological score to a screening test for cognitive impairment (Montreal Cognitive Assessment test - MoCA test <17.54; Conti et al., 2015) for PD patients.

### Trial Interventions

The Trial protocol provides for the random allocation of participants to two different types of intensive rehabilitation treatment: the SIDERA^B treatment (S) and the Usual Care treatment (U) according to a single-blind, crossover study approach.

### The SIDERA^B treatment (S)

Participants will receive 5 sessions/week (30-40 minutes/day) of an individualized, home-based telerehabilitation program delivered through the SIDERA^B platform, for the length of 3 months (for CHF patients) or 4 months (for COPD and PD patients). Each session will bring together motor *telerehabilitation* activities (Endurance Training module plus Resistance Training module or Neuromotor Training module) with *tele-monitoring* of vital parameters and health status, and motivational support for well-being (*tele-engagement*).

Participants will receive a home-based technological kit including a tablet delivering an individualized daily rehabilitation program and several medical devices for vital-signs monitoring (e.g., activity tracker, blood pressure monitor, balance, bicycle ergometer - *Davenbike* and pulse oximeter).

#### Tele-rehabilitation

This component will include a cardiovascular rehabilitation protocol (endurance training and resistance training modules) for COPD and CHF and a neuromotor rehabilitation protocol (endurance training and dance training modules) for PD. The training modules are digitalized (app) with the aim to foster internal adaptive loops for self-management in an asynchronous modality, improving the quality of care at home. In more detail, each training module will be described as follows:

– the *Endurance Training App* provides for aerobic exercises performed with the *Davenbike* bicycle ergometer in safety (i.e., sitting position) for the enhancement of cardio-pulmonary strength in all clinical conditions considered in the trial. This training, prescribed 3 times/week, last about 30 minutes and includes three main phases: “Warm-up” (lasting 5 minutes), “Exercise” (for about 20 minutes) and “Cool-down” (lasting 5 minutes). This app integrates the home training with clinical response parameters collected by an activity tracker for the self-monitoring of the heart rate in real-time: during the aerobic exercise, the tablet communicates changes in heart rate to the patient, who must increase or decrease the intensity of the physical effort according to the heart rate range set by the clinician on the technological platform. The chosen workload during the Endurance training will reflect the individual effort tolerance with regard to (1) perceived exertion according to the Borg scale (Borg, 1998) and (2) the heart rate range established individually for each patient according to the under-strain baseline evaluation. The Endurance training app is also gamified with a variety of multimedia digital contents which allow participants to every day explore, by bicycle, different places in the world while performing their rehabilitation, enhancing the level of patient participation and engagement.
– the *Resistance Training App* allows for patient-tailored muscle-strengthening through specific multimedia digital contents. This training, prescribed twice a week, integrates the monitoring of fatigue and dyspnea levels recorded by validated clinical scales (BORG RPE and CR10 scales (Borg, 1998). Each session lasts about 30 minutes and involves different exercises for patients with CHF and COPD patients, delivered on the tablet. Every multimedia activity starts with a brief explanatory training conducted by the physiotherapist showing the correct execution of the exercises. Then, the subject performs the exercises while observing the physiotherapist, following his own rhythm. The chosen workload during the Resistance training reflects the individual effort tolerance concerning (1) the perceived exertion according to the Borg scale, (2) the performance of the subject, and (3) the recovery time needed by the patient. Interestingly, the therapist could monitor these parameters over time through the SIDERA^B platform and tailor the type and intensity of the exercises according to the patient’s needs and performance.
– the *Neuromotor Training App* (Dance therapy) have been developed specifically for patients with PD to enhance movement, coordination and balance while promoting cognitive and social aspects. In SIDERA^B, this training is prescribed twice a week and delivered through specific multimedia contents that involve four different dance styles performed by a professional dancer. Each style includes 8 sessions lasting about 50-60 minutes which combine steps and sequences of movement patterns of increasing complexity in complete safety (i.e., goal-directed repetitive practice). To facilitate skill learning action observation strategies are implemented and complex patterns of movements have been unpacked into simpler components (with and without music) before practicing the whole choreography.

Overall, participants will freely decide at what times to perform the exercises, in relation to their preferences and needs. Data about rehabilitation sessions (e.g., if and how the exercises will be performed) will be automatically recorded on the SIDERA^B platform.

#### Tele-monitoring

This component will provide for the monitoring of vital parameters and adherence to drug treatment through different medical devices integrated into the SIDERA^B platform and supplied at the patient’s home according to the clinical condition: CHF home kit will include a blood pressure monitor, a balance and a pulse oximeter, COPD patients will receive a blood pressure monitor, a pulse oximeter and a technological device for the monitoring of compliance to the inhalation therapy, and PD patients will be provided with the blood pressure monitor and a “Parkinson’s diary” in order to monitor the presence/absence of abnormal movements or walking difficulties on a daily basis. Data from medical devices will be transmitted to the SIDERA^B platform by tablet for being seen at regular intervals (once a week) by healthcare professionals.

#### Tele-engagement

The core idea behind the component of tele-engagement for well-being (delivered through “The Living Book” app) is that tele-rehabilitation requires the appropriation of novel at-home care routines and that the appropriation of these routines is mediated by the fostering of well-being resources. Thus, engagement is meant as an appropriation process (rather than outcome) and is an integral part of the tele-rehabilitation program. The rationale is that by providing the patient with experiential affordances that tap into specific well-being resources, tele-rehabilitation becomes a holistic sense-making process that facilitates the management of uncertainty in the face of the day-to-day challenges of living with a chronic illness. “The Living Book” is an applied game, that is a user-centric experiential tool that borrows elements from traditional videogames but is designed as an intervention tool for a specific real-world problem. The core of the narrative design is a metaphor related to living with a chronic condition and its amelioration through specific psychological well-being resources. The book is organized as a sequence of stories, and patients have the opportunity to read and interact with one new story each day. Through their choices, they shape how the narrative unfolds. The storyline and gameplay follow the transformation of the main characters as they reflect upon and master the different well-being resources: autonomy, mutuality, cooperation, and purpose. The stories have been modelled to express and invoke different aspects of those resources, encoded in the emotional experiences and development of the characters. The game interface is that of an interactive book which allows the navigation within and between stories through appropriately positioned user interface elements. A peculiar feature of The Living Book is that the journey of characters in the story is shown on a map covered by the Night’s Vale (a metaphor for chronic disabilities’ impact on movement). The Night’s Vale can be gradually revealed only through the use of a crystal. The crystal is directly connected to the SIDERA^B application servers and receives gameplay data, concerning the level of completion of the rehabilitation exercises assigned to each patient. Completing the assigned daily exercises generates more energy in the crystal and allows patients to reveal more of the game map. Through this feature, adherence to the telerehabilitation activities is integrated into the narrative of the story, so that the progression in the story symbolically embeds the patient’s progression in the rehabilitation program. Rooted in The Living Book’s design and its technical implementation is a system of metrics intended to track patients’ behavior within the game and, through the use of these data, allow clinicians and care managers in making more informed decisions regarding patients’ treatment plans.

### The Usual Care treatment (U)

As the SIDERA^B treatment, the Usual Care treatment will require a commitment of about 30-40 minutes a day, for five days a week, for 3 months (patients with CHF) or 4 months (patients with COPD and PD). Participants will undergo a standard outpatients rehabilitation program at home through the use of a manual with conventional indications about the rehabilitative exercises (i.e., aerobic activities and strengthening) and the vital parameters to be monitored (e.g., measurement of pressure, oxygenation). Participants will also be asked to complete a daily paper diary reporting self-monitoring data collected. A phone number of the Clinical Center will be also included in the rehabilitation manual to contact the clinicians if needed.

### Outcome Measures

Participants will undergo an extensive evaluation at the baseline (T1) and at each time-point of evaluation (T2, T3, T4).

### Primary outcome measures

1. Change in activation of patients measured by the Patient Activation Measure scale (**PAM13**: Hibbard et al., 2004).
2. Change in activity and participation measured by the WHO Disability Assessment Schedule 2.0 **(WHODAS 2.0**: World Health Organization, 2004; Federici et al., 2009).

### Secondary outcome measures

1. Change in patient’s self-rated health measured by the EuroQol five dimensions (5D) and five levels (5L) (**EQ-5D-5L**: The EuroQol Group, 1990; Janssen et al., 2013).
2. Change in care-relationship measured by the Mutuality Scale (**MS**: Archbold et al., 1990; Pucciarelli et al., 2016; Dellafiore et al., 2018).
3. Changes in scores of environmental mastery measured by the Psychological Well-Being scale (**PWB**: Ryff et al., 1996; Ruini et al., 2003).
4. Change in Dispositional Optimism measured by the Life Orientation Test-Revised (**LOT-R**: Scheier et al., 1994).
5. Change in resilience measured by the Connor-Davidson Resilience Scale (**CD-RISC** 10-item version: Campbell-Sills& Stein, 2007).
6. Changes in affect measured by the Positive-Negative Affect Schedule (**PANAS**: Watson et al, 1988).
7. Change in health status scores and Qol measured by:
  a. the Kansas City Cardiomyopathy Questionnaire (**KCCQ**: Green et al., 2000; Miani et al., 2003) only in patients with CHF;
  b. the Parkinsons’ Disease Questionnaire (**PDQ**: Jenkinson, 1997; Galeoto et al., 2018) only in PD patients;
  c. the St George’s Respiratory Questionnaire (**SGRQ**: Jones et al., 1991) only in patients with COPD.
8. Maintenance or improvement of Aerobic Capacity and Gait using the 6’ Minute Walking Test (**6MWT**: https://www.sralab.org/rehabilitation-measures/6-minute-walk-test) in all clinical conditions.
9. Changes in Balance, Functional Mobility and Strength measured by the 30 Second Sit to Stand Test (**30CST**: https://www.sralab.org/rehabilitation-measures/30-second-sit-stand-test) only in patients with CHF.
10. Change in motor functionality measured by the Movement Disorder Society - Unified Parkinson’s Disease Rating Scale (**MDS-UPDRS**: Goetz et al., 2004), part-III, only in the PD group.
11. Change in the general cognitive domain and executive functioning measured by the Montreal Cognitive Assessment test (**MoCA**: Conti et al., 2015) and the Picture Interpretation test 360 (**PIT360**: Serino et al, 2017) only in PD patients.
12. Maintenance of cardiovascular performance measured by echocardiography and measure of Ejection Fraction (**EF**) using echocardiography as surrogate markers (only in CHF patients).
13. Maintenance of the Forced expiratory volume in the 1st second (**FEV1%**) measured by spirometry: the measure of FEV1% using spirometry as a surrogate marker (only in COPD patients).
14. Maintenance of structural brain indices and improvement of functional and metabolic parameters measured by Magnetic Resonance Imaging (**MRI**) investigation (only in PD – facultative data collection).

### Data collection

Demographic characteristics will be collected at the baseline evaluation (T1); data from participation measures and clinical/functional measures (primary and secondary outcomes) will be collected by blinded examiners/assessors at the baseline (T1) and at each time-point of evaluation (T2, T3, T4); finally, data on adherence to the rehabilitative program, performance levels at exercises and vitals-signs (e.g., heart rate, weight, oximetry, blood pressure) will be automatically collected through the SIDERA^B platform. Moreover, adverse events related to intervention throughout the study duration will be recorded. Finally, additional data useful for a technological evaluation will be collected with validated questionnaires, approaching with MAST technique (Kidholm, 2010), and following the EUnetHTA Core Model (2016) principles, developing organizational, legal, and equity impacts. Moreover, an economic and sustainability evaluation (Mauskopf et al., 2007) will be performed, as well as technological acceptability using the Technology Acceptance Model (TAM, Davis, 1989; Venkatesh & Davis, 2000) and *ad hoc* case report forms.

### Statistical analysis

All statistical analyses will respect the within-participant nature of the comparisons. Descriptive statistics of the sample will include frequencies, median and interquartile range (IQR) for categorical variables and Mean and Standard Deviation (SD) for continuous measures. The assumption of normality will be checked by the Shapiro-Wilk test for continuous variables. We will investigate statistically significant changes in primary and secondary outcome measures according to the Consolidated Standards of Reporting Trials (CONSORT) guidelines extended to randomized crossover trials. Statistically significant changes will be investigated using t-tests on within-participant differences, and analysis of variance with participant, period, and treatment effects. Period and carry-over effects will be modelled in the statistical plan.

## Discussion

In the last 30 years, the demand for rehabilitation services for NCSs has increased double and the need for rehabilitation currently accounts for about 2.41 billion individuals (Cieza et al., 2020). This worldwide phenomenon urgently requires new solutions to reach all people in need. In this framework, telemedicine and telerehabilitation are ideal candidates but just like a new drug, these digital medical solutions will require rigorous validation adopting RCT designs (Topol 2019b). The present crossover RCT is designed to test the effectiveness of an innovative digital, home-based rehabilitation program, compared to conventional face-to-face rehabilitation, in ensuring the continuity of care among patients with NCDs (CHF, COPD, and PD).

The SIDERA^B platform has been implemented and developed to improve new rehabilitative practices enabling a double-loop communication exchange between the hospital and the patient’s home. The asynchronous approach ensured by the digital platform allows for overcoming the limits imposed by traditional face-to-face interventions, improving the quality of long-term care at distance and increasing accessibility to a broader target (Dorsey et al., 2016). Through the SIDERA^B platform, rehabilitation sessions can be tailored to patients’ needs, and the therapist can monitor at any time the progression of the rehabilitation program. According to the daily outcomes recorded in the digital platform, healthcare professionals can track, for each patient and even in real-time, vital parameters, performance data, and adherence to treatment, adapting the prescription in a tailored and disease-specific manner. Moreover, the integration of digital contents and technological devices for telerehabilitation, telemonitoring and tele-engagement is promising for the improvement of the accuracy and efficiency of medical practice and for the promotion of a high degree of patient empowerment (Topol, 2019a).

To test the effectiveness of the SIDERA^B program, we will investigate after-treatment changes in Patient-Relevant Structural and Procedural Effects and Medical Benefits outcomes through a crossover design comparing patients in the SU Group and in the USU Group. Incorporating a crossover into a randomized study represents a powerful method to test the clinical effectiveness of interventions for important health outcomes. In addition, crossover study design can be advantageous in terms of a better balance between arms and increased statistical efficiency (Reich et al., 2014).

We expect that SIDERA^B may have several positive healthcare effects on health outcomes, such as more effective management of disease minimization of limitations in everyday functioning due to chronic health conditions. In addition, we look forward to an increase in activation and participation of patients and their caregivers in the healthcare process, due to the close attention on motivational aspects, psychological well-being and engagement.

In conclusion, SIDERA^B could represent a promising, innovative and sustainable digital solution able to support the actual migration of care from hospitals and clinics to the patient’s home, for the optimal long-term management of chronic conditions.

## Data Availability

All data produced in the present study are available upon reasonable request to the authors

https://clinicaltrials.gov/ct2/show/NCT04041193

## Declarations

### Ethics approval and consent to participate

The study was approved by the Ethical Committee of IRCCS Don Carlo Gnocchi Foundation and was registered as a clinical trial on clinicaltrials.gov (identifier NCT04041193). Prospective participants will be fully informed of the aims and procedures of the project. A reporting procedure will be in place to ensure that any serious adverse events are reported to the Chief Investigator. Informed written consent will be obtained from all participants before the study initiation.

## Acknowledgements

We acknowledge the support of all the staff of the SIDERA^B Consortium. The SIDERA^B Consortium consisted of: ab medica S.p.A. (Cerro Maggiore, Milano); IRCCS Fondazione Don Carlo Gnocchi ONLUS (Milano); Dipartimento di Scienze Umane per la Formazione “Riccardo Massa” e Dipartimento di Informatica, Sistemistica e Comunicazione, Università degli Studi di Milano-Bicocca (Milano); Università Carlo Cattaneo-LIUC (Castellanza, VA); Politecnico di Milano (Milano); Grifo Multimedia S.r.l. (Ruvo di Puglia, BA); Tenacta Group S.p.A. (Azzano San Paolo, BG); Amiko S.r.l. (Milano).

## Authors’ contributions

Developed the original concept of the trial (FBA, FR, OR, EF, DC) developed the design and methodology (FBA, MN, PIB, MT), developed the analysis plan (FBO, OR), adapted the trial proposal as a protocol paper (FR, VB, PIB, MT), did manuscript writing (FR, FBO, EG). All authors reviewed and commented on drafts of the protocol and paper. All authors read and approved the final manuscript.

